# Distinct Evolutionary Trajectories in Early Lung Adenocarcinoma: Age-Related Pathway with Epidermal Growth Factor Receptor–Associated Genome Doubling and Smoking-Driven Pathway

**DOI:** 10.64898/2026.07.21.26358543

**Authors:** Akiteru Goto, Hirofumi Nakaoka, Makoto Yoshida, Kei Koyama, Ken Miyabe, Jian Zhou, Michinobu Umakoshi, Shinogu Takashima, Kazuhiro Imai, Yoshihiro Minamiya, Keiko Nishikawa, Daisuke Matsubara, Ituro Inoue, Haruhiko Sugimura, Yuichi Ishikawa

**Author notes:** **Corresponding Authors** Akiteru Goto, MD, PhD, Department of Quantitative Pathology, Akita University Graduate School of Medicine, 1-1-1 Hondo, Akita 010-8543, Japan, Hirofumi Nakaoka, PhD, Department of Biomedical Data Science, Kagoshima University Graduate School of Medical and Dental Sciences, 8-35-1 Sakuragaoka, Kagoshima 890-8544, Japan. These authors contributed equally to this work (Akiteru Goto, Hirofumi Nakaoka). These authors jointly supervised this work (Haruhiko Sugimura, Yuichi Ishikawa).

## Abstract

Lung adenocarcinoma a progress from preinvasive lesions to invasive cancer; however, early evolutionary events in adenocarcinoma *in situ* (AIS) and minimally invasive adenocarcinoma (MIA) remain poorly defined, particularly in East Asian populations enriched for *EGFR* mutations. We performed whole-exome sequencing on 67 Japanese patients (38 AIS, 29 MIA), including multiregion sampling in 13 cases to identify *EGFR* as predominant driver (61.2%), followed by *RBM10* (19.4%) and *TP53* (9.0%). Two evolutionary trajectories emerged: age-related and smoking-driven pathways. In the former, *EGFR* mutant tumors frequently exhibited early whole genome doubling (WGD) (24.4%) with clock like signature. The smoking driven pathway, typically involving *KRAS* mutations, displayed a tobacco associated signature. Multiregion sequencing revealed that driver mutations *(EGFR*, *KRAS*, and *MET*) were shared trunk events across *in situ* and invasive regions, while secondary alterations arose subclonally. This study defines the genomic evolution of early lung adenocarcinoma in Japanese patients, identifying two evolutionary trajectories: an age-related pathway with *EGFR-*linked genome doubling and a smoking-driven pathway involving *KRAS* mutations. These findings elucidate mechanisms underlying progression from preinvasive lesions to invasive cancer in Asian populations.

## INTRODUCTION

Lung cancer remains one of the world’s most prevalent malignancies and the leading cause of cancer-related mortality globally (1). Among lung cancers, lung adenocarcinoma is the most common histological subtype and continues to increase in incidence (2). Pathological and genetic evidence supports a multistep progression model in which the precursor lesion atypical adenomatous hyperplasia (AAH) progresses to adenocarcinoma *in situ* (AIS), minimally invasive adenocarcinoma (MIA), and ultimately invasive nonmucinous adenocarcinoma (IA) (2,3). Although large-scale sequencing projects conducted by The Cancer Genome Atlas Research Network and the Pan-Cancer Analysis of Whole Genomes have defined the genomic landscape of lung adenocarcinoma, including somatic single nucleotide variants (SNVs), short insertions and deletions (indels), somatic copy number alterations (SCNAs), and structural variants, most analyses have focused on advanced disease (4,5). In contrast, genetic alterations in early-stage lung adenocarcinoma, particularly AIS and MIA, remain incompletely characterized, and it is unclear whether findings from advanced tumors can be extrapolated to early lesions. Therefore, the proposed sequence from AAH through AIS and MIA to IA requires further investigation, with emphasis on early-stage disease.

Because multistep lung adenocarcinoma progression is driven by repeated clonal expansions acquiring advantageous mutations (6), differences between early and advanced tumors may reveal stage-specific molecular drivers. Consequently, genomic studies focusing of early lung adenocarcinoma are garnering increasing attention (7). Prior work, including our own, has identified mutations in epidermal growth factor receptor (*EGFR*) and Kirsten rat sarcoma viral oncogene homolog (*KRAS*) in AAH, AIS, and MIA as early oncogenic events (8). Moreover, total mutational burdens increase stepwise from AAH through AIS and MIA to invasive lung adenocarcinoma, accompanied by rising clonal and subclonal mutational burdens, SCNAs, and allelic imbalance (7). These findings support a general multistep model of lung adenocarcinoma development.

Although lung cancer is strongly linked to tobacco exposure, its incidence in never-smokers remains a major global health concern, particularly among East Asian females (2). Genomic studies have associated *KRAS* mutations with smokers and *EGFR* mutations with never-smokers. Beyond mutational burden, mutational signature analysis provides a record of tumorigenic processes. Comprehensive catalogs link specific single-base substitution (SBS) patterns to distinct etiologies, including DNA-repair defects and environmental exposures (9). Tobacco exposure typically produces a C > A transversion signature (SBS4), whereas the origins of lung adenocarcinoma in never-smokers are less well defined. Genomic classifications suggest that lung cancer in never-smokers (LCIN) follows distinct evolutionary routes driven by endogenous processes, such as aging or apolipoprotein B mRNA editing enzyme catalytic polypeptide-like (APOBEC) activity (10). However, how these processes shape genomic alterations in AIS and MIA remains unclear. Therefore, studying these precursor lesions is essential to determine whether signatures observed in advanced never-smoker lung adenocarcinoma arise early in tumorigenesis.

Genomic heterogeneity drives tumor evolution, making multiregion sampling critical for characterizing intratumoral clones (11,12). Although multiregion sequencing has revealed extensive intratumoral heterogeneity in advanced cancers, its extent in AIS and MIA is incompletely documented. Conventional bulk sequencing cannot fully reconstruct evolutionary histories, as it may miss spatially distinct subclonal mutations that promote invasion. Therefore, we applied multiregion sampling to distinguish “trunk” mutations, i.e., early events shared by all cells, from “branch” mutations, restricted to subclones. Determining whether invasion drivers arise early or during subclonal diversification is crucial for understanding early lung adenocarcinoma progression.

Specifically, we performed whole-exome sequencing (WES) on 103 samples from 67 Japanese patients with early lung adenocarcinoma and characterized the genomic features of early lung adenocarcinoma, including driver mutations, somatic SNV and SCNA burdens, and mutational signatures. Our findings show that mutational burden in early lung adenocarcinoma reflects an interplay between driver gene status and patients’ smoking history.

## MATERIALS AND METHODS

### Lung AIS and MIA cases

We analyzed 67 early lung adenocarcinoma cases: 38 AIS and 29 MIA. Patient demographics are summarized in Supplementary Table 1. Clinicopathological data, including age, sex, and cumulative SI, were obtained from clinical records. Archival pathological samples from the 67 cases were retrieved from surgical specimens obtained at Akita University Hospital (Akita, Japan) during 2014–2022. None of the patients had received preoperative chemotherapy. Histopathological diagnoses were assigned according to the World Health Organization classification system, and all tumors were confirmed as AIS or MIA (2). Representative slides were scanned at 20× magnification using a pathology digital imaging system (Nanozoomer virtual slide system, Hamamatsu Photonics, Shizuoka, Japan). Whole-slide images were independently evaluated by three pulmonary pathologists (A.G., D.M., and Y.I.) to confirm diagnostic agreement. Ethical approval was obtained from Akita University, Faculty of Medicine, Ethics Committee (Reference No. 1241), and written informed consent was obtained from each patient.

### Whole-tumor sampling

DNA was extracted from macrodissected formalin-fixed paraffin-embedded (FFPE) 40–50-μm-thick tumor tissue sections (4–5 serial sections with 10-μm thickness) from each case using FormaPure XL Total (BECKMAN COULTER, Tokyo, Japan). DNA was extracted from background lung tissue in each case in a similar manner.

### Multiple location sampling

In three AIS and ten MIA cases, three rectangular regions were individually macrodissected from FFPE tumor tissues along with one region from adjacent nontumorous lung tissue. Sampling areas ranged from 9 to 16 mm^2^ and were arranged from the tumor center to the periphery. In MIA cases, central regions corresponded to invasive areas and peripheral regions to noninvasive (*in situ*) components. DNA extraction followed the same protocol used for whole-tumor sampling.

### WES

WES was performed as previously described (13). Extracted DNA was repaired using NEBNext FFPE DNA-Repair Mix (New England Biolabs) and fragmented with a NEBNext dsDNA Fragmentase (New England Biolabs). Libraries were prepared using the NEBNext Ultra II DNA Library Prep Kit (New England Biolabs Japan, Tokyo, Japan), and target enrichment was conducted with the xGen Exome Research Panel v2 (Integrated DNA Technologies, Tokyo, Japan). Library sequencing was performed on an Illumina NovaSeq 6000 platform using a 2 × 150-bp paired-end configuration (Illumina, San Diego, CA, USA).

### Quality control of sequencing data and variant calling

Detailed bioinformatics pipelines for sequencing data quality control and variant calling were described previously (13,14) and are provided in the supplementary notes.

### Mutational signature analysis

Somatic SNVs with mutant allele frequency (MAF) ≥0.10 from all 67 patients were used for mutational signature analysis. Tumors were classified into five groups according to driver gene mutation and smoking statuses: *EGFR*-NS, *EGFR*-SM, non-*EGFR* CAG-NS, non-*EGFR* CAG-SM, and No CAG-NS. In each group, somatic SNVs were classified into 96 mutation classes defined by six pyrimidine substitutions (C > A, C > G, C > T, T > A, T > C, and T > G) and their flanking 5′ and 3′ bases.

In mutational signature analysis, the 96-mutation catalog was fitted to predefined signatures (15,16) rather than *de novo* extracted signatures because mutation counts were insufficient for reliable *de novo* analysis. COSMIC mutational signatures version 3 served as the reference dataset (9). We selected 11 SBS signatures (SBS1, SBS2, SBS4, SBS5, SBS8, SBS13, SBS17a, SBS17b, SBS18, SBS45, and SBS92) with activities estimated to be present in lung adenocarcinoma based on the COSMIC mutational signatures. Signature fitting was achieved using sigfit (bioRxiv 2018.10.22.448969). Four Markov chains were run for 50,000 iterations, including a burn-in of 25,000 iterations. Highest posterior density (HPD) intervals were estimated for each SBS signature. Signatures with a 90% lower HPD bound exceeding 0.01 (default value) were considered significantly active.

### Detection of SCNAs

SCNAs were detected using FACETS (17) based on total read depth and allelic imbalance in tumor and matched background lung samples. Germline polymorphic sites were obtained from variant call format (VCF) files generated by the 1000 Genomes Project (18).

### Intratumor genomic heterogeneity

To evaluate clonal relationships among spatially distinct lesions, we compiled MAF profiles for all mutation sites across AIS, MIA and adjacent nontumorous lung samples from each patient. Reference and mutant allele reads were counted using SAMtools mpileup (arXiv:1303.3997). Only high-confidence reads with mapping quality >30 were used, and allele-specific counts were measured using only base calls with base quality >20 at mutation sites. The MAF profiles of informative mutation sites were analyzed using hierarchical clustering analysis in the superheat R package (19).

### Statistical analyses

Associations between gene mutation status and clinicopathological variables were assessed using Fisher’s exact test. Differences in mutation frequencies between early lung adenocarcinoma in the current study and advanced lung adenocarcinoma in the IntOGen database were also assessed using Fisher’s exact test. Co-occurrence and mutual exclusivity analyses of cancer-associated gene mutations were similarly performed using this test. All Fisher’s exact tests were implemented using the fisher.test function in the stats R package. Associations between clinicopathological variables and mutational burden were evaluated using the exact Wilcoxon rank-sum test for binary categorical variables (exactRankSum R package), the Kruskal–Wallis rank-sum test for variables with three or more categories, and the Jonckheere–Terpstra trend test for ordered variables (DescTools R package). A two-sided *P*-value < 0.05 was considered statistically significant.

### Visualizations

Methods for data visualization are described in the supplementary notes.

### Data availability statement

The datasets generated in this study are available from the corresponding author upon reasonable request.

## RESULTS

### Mutational landscape of early lung adenocarcinoma

To identify somatic mutations in early lung adenocarcinoma, we performed WES on 38 AIS and 29 MIA cases (Supplementary Table 1). The mean sequencing depth and percentage of exome covered by ≥20 reads averaged 190.3 and 96.9%, respectively (Supplementary Table 2). The mutational landscape of early lung adenocarcinoma is shown in Figure 1a. The most frequently mutated gene was *EGFR* (41/67, 61.2%), followed by RNA binding motif protein 10 (*RBM10*: 13/67, 19.4%), tumor protein p53 (*TP53*: 6/67, 9.0%), *KRAS* (6/67, 9.0%), and LDL receptor-related protein 1B (*LRP1B*: 5/67, 7.5%). Associations between mutation status and clinicopathological features were also evaluated (Supplementary Table 3). *KRAS* mutations were overrepresented in moderate-to-heavy smokers (P = 3.9 × 10 □□) and males (P = 4.8 × 10□³), whereas associations for *LRP1B* and *RBM10* mutations with moderate-to-heavy smoking were nominally significant (P < 0.05).

**Figure 1.**
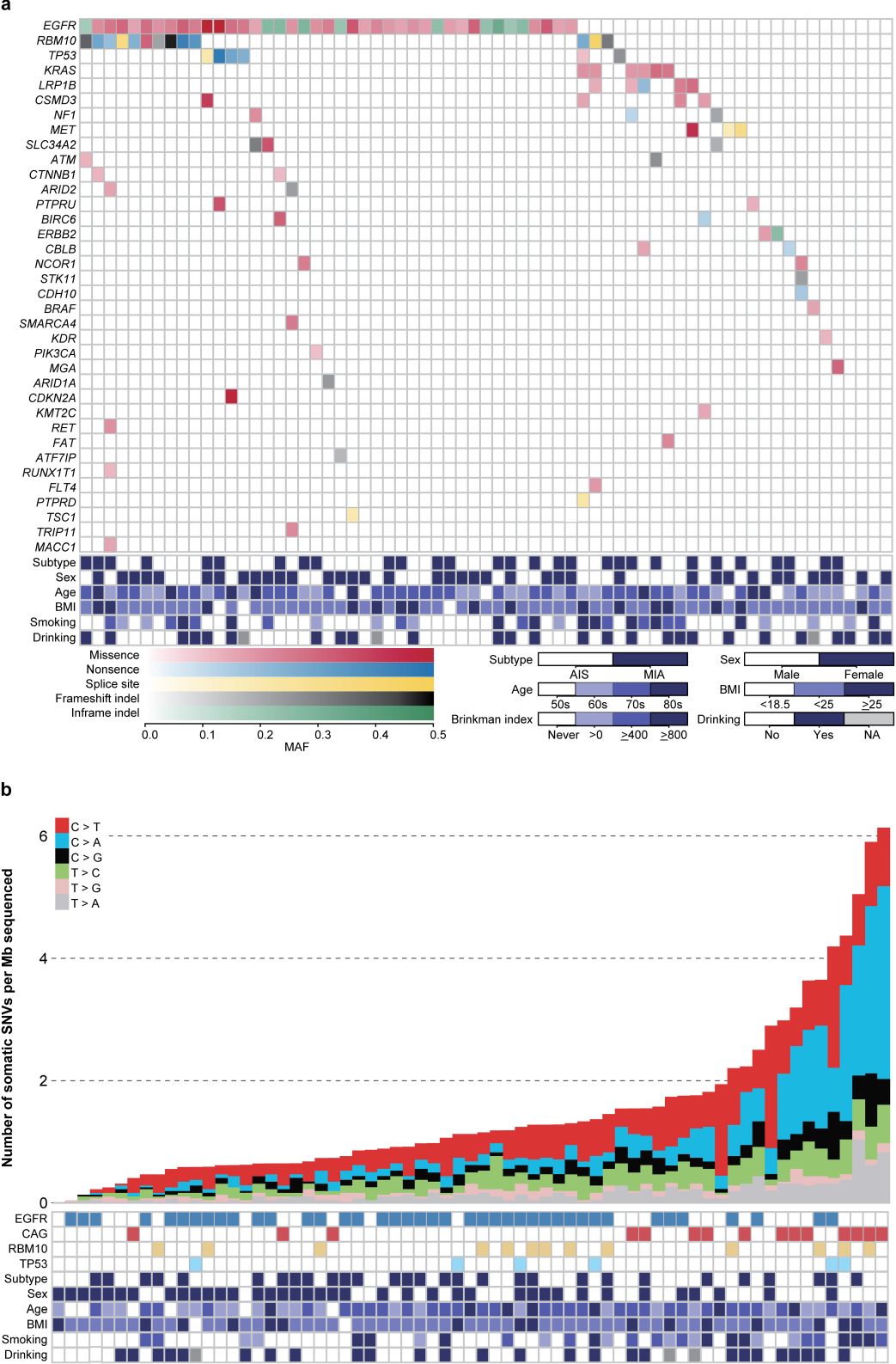
Mutational landscape of early lung adenocarcinoma. (A) Heatmap representing the mutation status of recurrently altered cancer-associated genes in AIS and MIA samples. Mutations are color-coded as missense SNVs (red), nonsense SNVs (blue), splice-site SNVs (yellow), in-frame indels (green), and frameshift indels (black). Color intensity indicates the MAF of each somatic mutation. (B) Bar charts showing somatic SNV burdens stratified by the six pyrimidine substitutions (C > T, C > A, C > G, T > C, T > G, and T > A). Burden is defined as the number of SNVs per megabase of sequenced genome. The lower heatmap summarizes gene mutation status and clinicopathological characteristics. CAG indicates mutations in cancer-associated genes, including *KRAS*, *LRP1B*, *NF1*, *MET*, *ERBB2*, *STK11*, and *BRAF*.

We mapped mutations to encoded protein domains, finding that *EGFR* mutations clustered at hotspots, including missense variants (p. L858R, p. L861Q) and in-frame exon 19 deletions (p. E746_A750del, p. L747_T751del) (Figure 2a). Notably, in-frame exon 20 insertions (p. A767_V769dup, p. D770_N771insG), typically resistant to EGFR tyrosine kinase inhibitors (20), were also detected. *RBM10*, a regulator of alternative splicing for multiple target genes (e.g., NUMB endocytic adaptor protein and eukaryotic translation initiation factor 4H) (21), harbored diverse inactivating mutations, consistent with a tumor suppressor role in early lung adenocarcinoma (Figure 2b). *TP53* showed missense mutations in its DNA binding domain along with other inactivating variants (Figure 2c). *KRAS* mutations occurred at codons 12 and 61, with all codon 12 variants being C > A transversions. MET proto-oncogene, receptor tyrosine kinase (*MET*) mutations were enriched at the exon 14 splice site, consistent with oncogenic activation (Supplementary Figure 1) (22). Compared with invasive lung adenocarcinoma in the IntOGen database (23), *EGFR* and *RBM10* mutations were more frequent in early lung adenocarcinoma, whereas *TP53*, *KRAS*, and *KEAP1* mutations were less common (Figure 2d, Supplementary Table 4).

**Figure 2.**
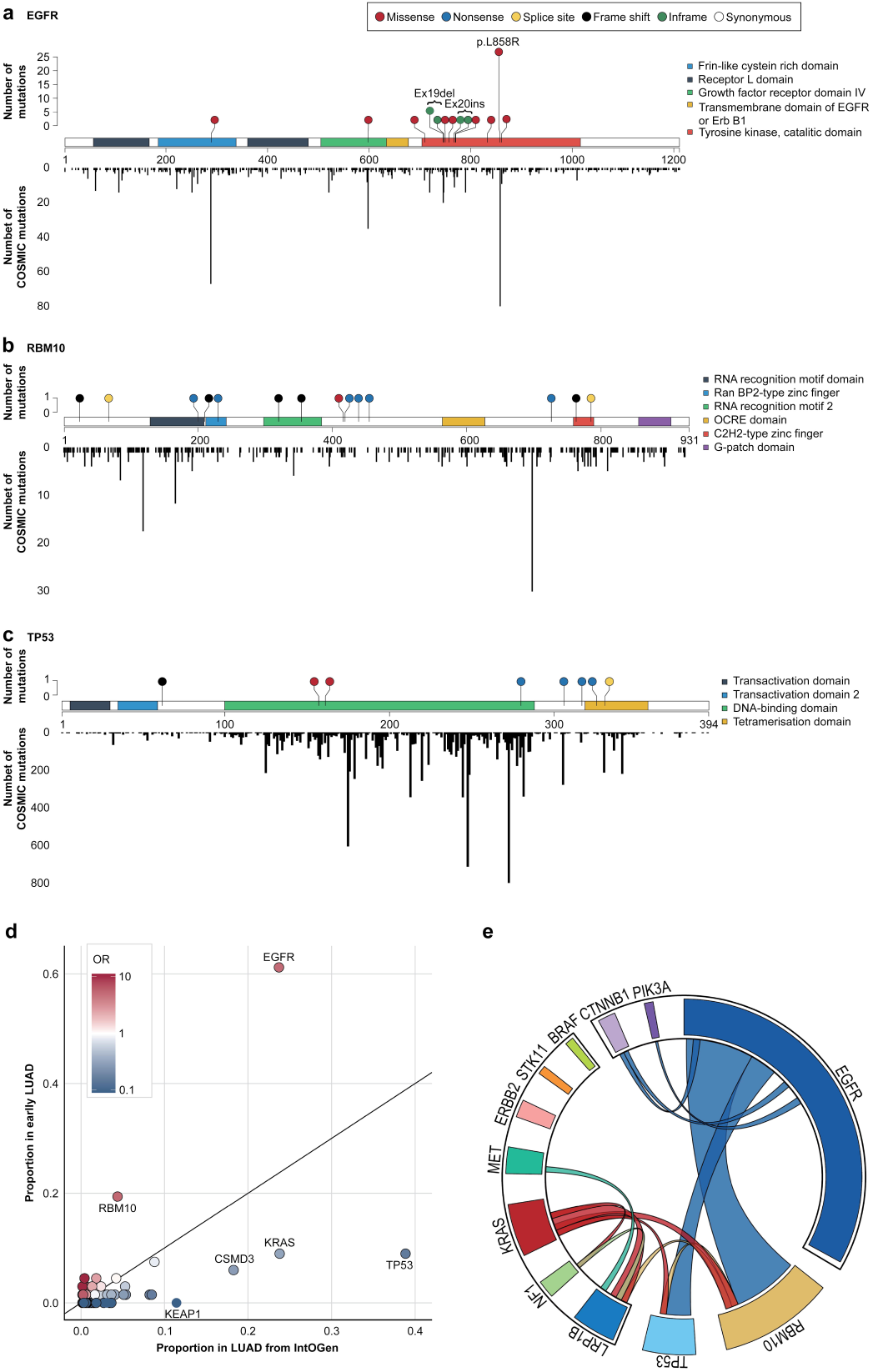
Localization, mutual exclusivity, and co-occurrence of cancer-associated gene mutations in early lung adenocarcinoma. (A–C) Lollipop plots showing distributions of somatic mutations in (A) *EGFR*, (B) *RBM10*, and (C) *TP53*. (D) Scatter plot comparing the mutation frequencies of cancer-associated genes between early lung adenocarcinoma in the current study and invasive lung adenocarcinoma from the IntOGen database. (E) Circos plot illustrating mutual exclusivity and co-occurrence patterns among cancer-associated gene mutations in early lung adenocarcinoma.

Next, we assessed the co-occurrence and mutual exclusivity of cancer-associated gene mutations in early lung adenocarcinoma (Figure 2e). Consistent with prior studies (24), *EGFR* and *KRAS* mutations were mutually exclusive (P = 9.3 × 10, Fisher’s exact test). Mutations in the oncogenes *MET* (n = 3), erb-b2 receptor tyrosine kinase 2 (*ERBB2*; n = 2), and Braf proto-oncogene (*BRAF*; n = 1) did not co-occur with *EGFR* or *KRAS* (P = 8.1 × 10 ¹). In contrast, *RBM10* or *TP53* mutations appeared in *EGFR*- and *KRAS*-mutant tumors, suggesting that *RBM10* and *TP53* mutations play co-operative roles with oncogenic drivers, such as *EGFR* and *KRAS*, in lung adenocarcinoma development.

### Mutational burden and signatures in early lung adenocarcinoma

Mutational burden varied widely across early lung adenocarcinoma cases and was largely driven by C > A transversions associated with smoking (Figure 1b). Consistent with previous research (25), *EGFR*-mutant tumors showed lower mutational burdens, whereas tumors with mutations in cancer-associated genes other than *EGFR* (non-*EGFR* CAG) had higher burdens. No significant difference in mutational burden was observed between AIS and MIA. Therefore, we stratified early lung adenocarcinoma samples into five groups based on mutation and smoking statuses; *EGFR*-mutant nonsmokers (*EGFR*-NS), *EGFR*-mutant smokers (*EGFR*-SM), non-*EGFR* CAG–mutant nonsmokers (non-*EGFR* CAG-NS), non-*EGFR* CAG–mutant smokers (non-*EGFR* CAG-SM), and tumors lacking known cancer-associated gene mutations (No CAG-NS) (Figure 3a). Analysis of 11 lung adenocarcinoma–associated SBS signatures from COSMIC (9) showed significant enrichment of clock-like signatures (SBS1, SBS5) (26), a smoking-related signature (SBS4), and APOBEC3-associated signatures (SBS2, SBS13) (Figure 3b). As expected, SBS4 mutations were increased in *EGFR*-SM and non-*EGFR* CAG-SM compared with *EGFR*-NS and non-*EGFR* CAG-NS, respectively. However, *EGFR*-SM tumors exhibited fewer SBS4 mutations relative to non-*EGFR* CAG-SM and a similar SBS4 mutational contribution compared with non-*EGFR* CAG-NS. SBS4 mutations predominated in non-*EGFR* CAG-SM. APOBEC3-related mutations were detected from the early stages of lung adenocarcinoma in all groups except non-*EGFR* CAG-SM. In No CAG-NS tumors, most mutations were attributed to SBS1, likely reflecting formalin fixation–induced artifacts (27). Reduced DNA quality may also explain the absence of detectable drivers in these samples. Notably, transcription strand asymmetry, with enrichment of SBS4-associated C > A mutations on the transcribed strand, was observed in the non-*EGFR* CAG-NS and non-*EGFR* CAG-SM groups (Figure 3c), reflecting interactions between DNA-damaging agents and DNA-repair mechanisms.

**Figure 3.**
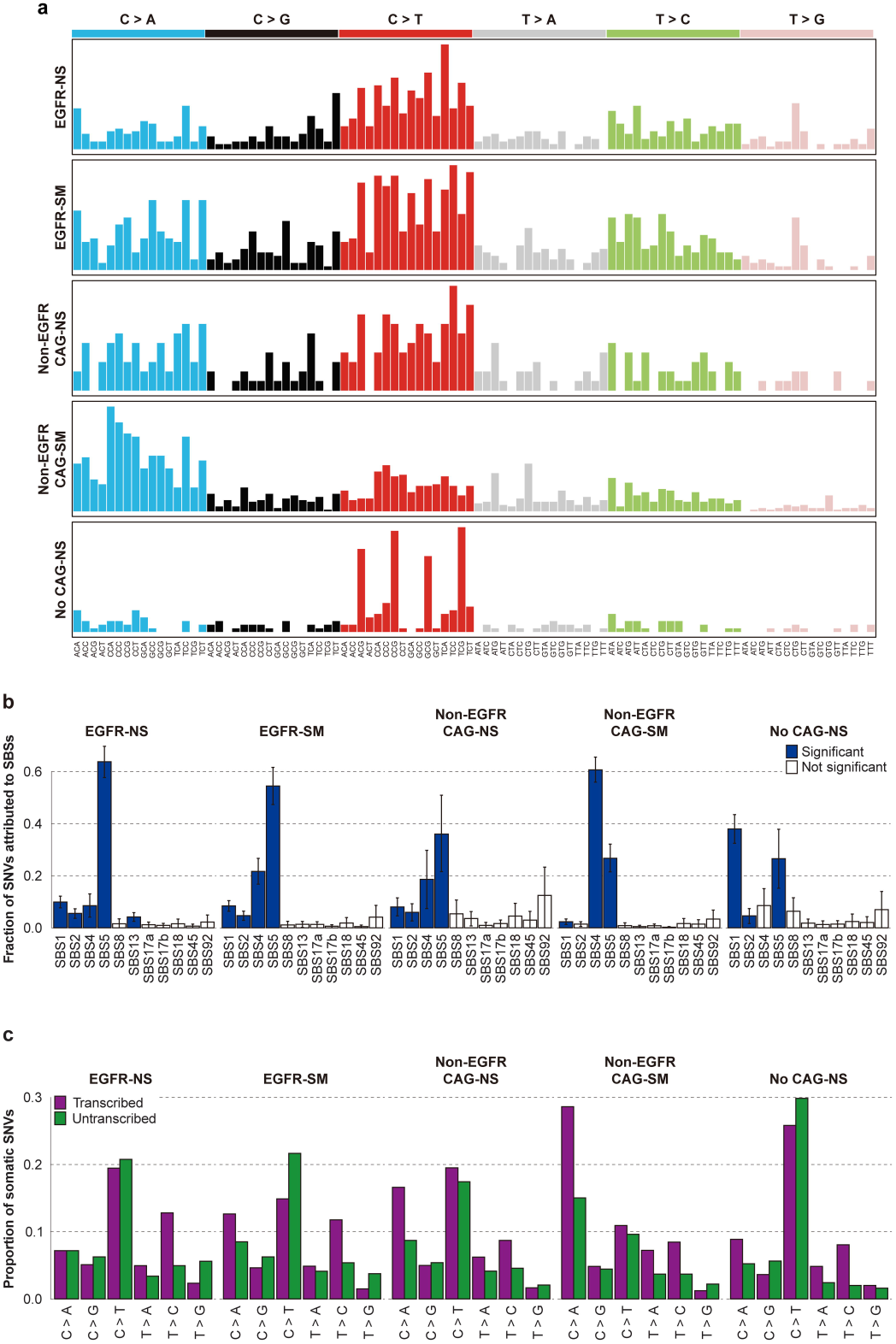
Mutational signatures and transcriptional strand bias. **(A)** Relative contributions of single-base substitution (SBS) types across molecular subgroups (n = 67): *EGFR*-mutant nonsmokers (*EGFR*-NS), *EGFR*-mutant smokers (*EGFR*-SM), non-*EGFR* CAG–mutant nonsmokers (non-*EGFR* CAG-NS), non-*EGFR* CAG–mutant smokers (non-*EGFR* CAG-SM), and tumors without known cancer-associated gene mutations (No CAG-NS). (B) Decomposition of mutational profiles into 11 COSMIC SBS signatures. Blue bars indicate significant enrichment, whereas white bars indicate nonsignificance. Error bars represent 95% confidence intervals. (C) Transcriptional strand asymmetry analysis showing proportions of somatic SNVs on transcribed (purple) versus untranscribed (green) strands, stratified by subgroup and substitution type.

### Associations between mutational burden and clinicopathological features

We examined associations between clinicopathological features and mutational burdens across the six substitution classes defined by the mutated pyrimidine base: C > A, C > G, C > T, T > A, T > C, and T > G (Supplementary Figures 2-1 and 2). Patient age positively correlated with mutational burden. Cumulative smoking exposure, measured via the smoking index (SI; cigarettes smoked per day × years smoked), showed strong positive correlations with C > A and T > C substitution burdens (P = 5.2 × 10□□and 2.8 × 10□□, respectively; Figure 4a). Tumors from males also exhibited higher mutational burdens, likely reflecting higher smoking prevalence among men. Furthermore, C > A and T > C mutation burdens differed significantly according to driver gene mutation status (P = 2.0 × 10□³ and 0.01, respectively; Figure 4b, Supplementary Figures 3 and 4). Histological subtypes (AIS vs. MIA), body mass index (BMI), and alcohol consumption showed no significant associations with mutational burden (P > 0.05; Supplementary Table 5).

**Figure 4.**
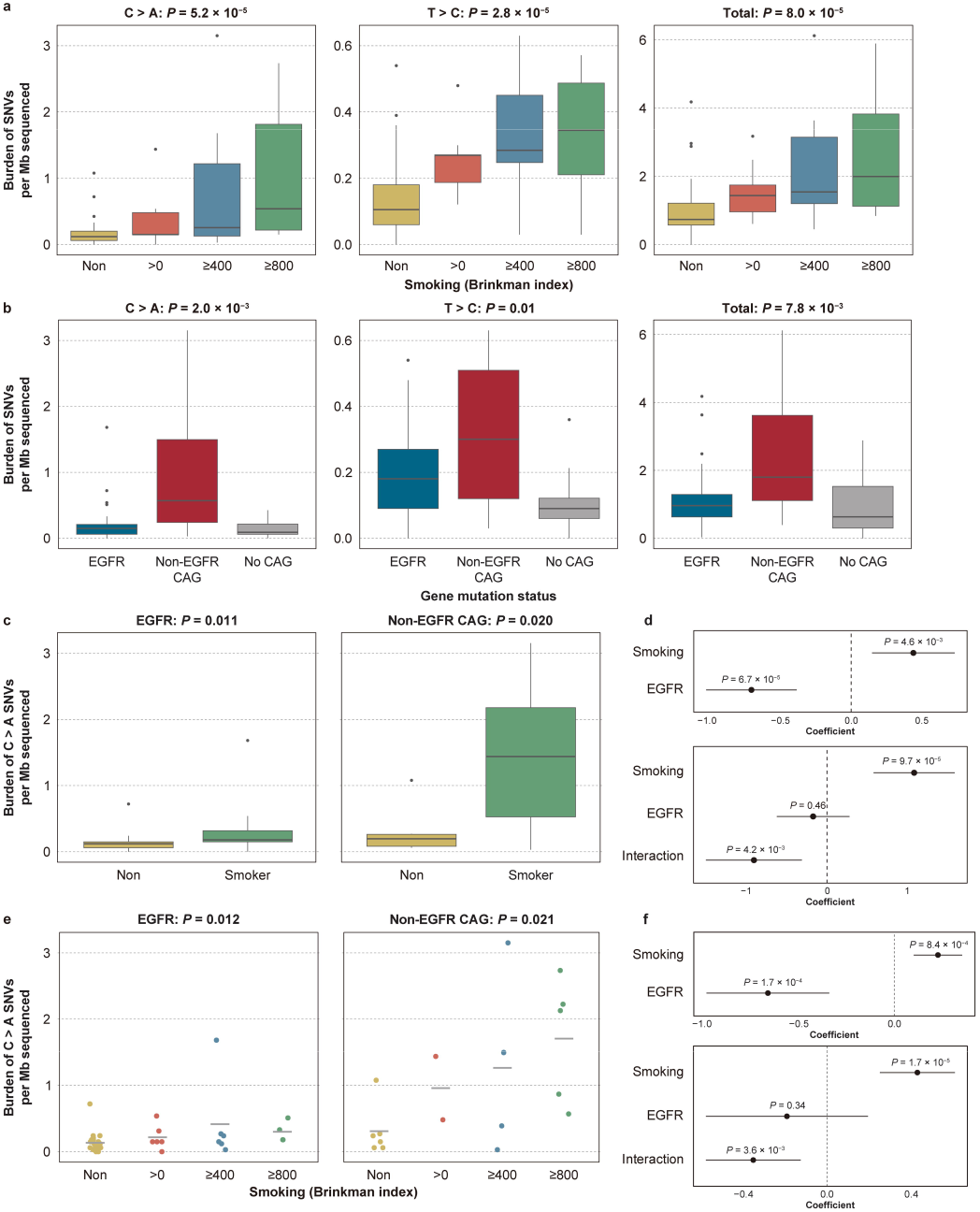
Interaction between smoking exposure and driver gene mutation status. (A) Box plots of C > A and T > C somatic substitutions per megabase relative to cumulative smoking exposure measured via the SI. P-values were calculated using the Jonckheere–Terpstra trend test. (B) Box plots of C > A and T > C substitutions stratified by driver gene mutation status (*EGFR*-mutant, n = 41; other CAG mutant, n = 17; no CAG, n = 9). P-values were determined using the Kruskal–Wallis test. (C–F) Comparisons of C > A mutation burden between nonsmokers and smokers (C) or according to cumulative smoking exposure measured via the SI (E), stratified by driver gene status. Coefficient plots from multivariable linear regression models show the independent and interaction effects of smoking and driver gene mutation status on C > A mutation burden, with smoking modeled as a binary variable (nonsmoker vs. smoker) (D) or cumulative exposure measured via the SI (F). Points indicate coefficients, and error bars indicate 95% confidence intervals. In all box plots, center lines represent medians, boxes indicate interquartile ranges, and whiskers extend to 1.5× the interquartile range.

Based on these findings, we hypothesized that mutational burden in early lung adenocarcinoma is shaped by interactions among smoking exposure and driver gene mutation status. Stratified analysis by driver gene mutation revealed that the increase in C > A mutation burden between smokers and nonsmokers was greater in non-*EGFR* CAG–mutant tumors than in *EGFR*-mutant tumors (Figure 4c). Multivariable linear regression, where smoking and driver gene mutation status were included as explanatory variables, confirmed that they were independently associated with C > A mutation burden. Addition of an interaction term between these factors showed a significant interaction effect (P = 4.2 × 10□³), indicating that smoking exerted significantly different effects on C > A mutation burden in *EGFR*-mutant versus non-*EGFR* CAG–mutant tumors (Figure 4d). This interaction remained significant when cumulative smoking exposure, modeled using SI, was considered (P = 3.6 × 10□³; Figure 4e, f). These findings indicate that cumulative smoking exposure exerts distinct, dose-dependent effects on C > A mutation burdens in *EGFR*-mutant and non-*EGFR* CAG–mutant tumors, with markedly greater effects in the latter. Similar interaction patterns were observed between smoking and driver gene mutation status for T > C mutation burden in both binary and dose-dependent smoking analyses, although these associations were suggestive rather than significant (P < 0.1; Supplementary Figures 3 and 4).

### SCNAs and whole-genome doubling events in early lung adenocarcinoma

Next, we examined SCNAs in early lung adenocarcinoma (Figure 5a). The proportion of the exome affected by SCNAs was greater in *EGFR*-mutant tumors than in non-*EGFR* CAG–mutant tumors (P = 5.1 × 10 ³). Allele-specific DNA copy number analysis using FACETS (17) showed that *EGFR*-mutant tumors had a larger proportion of the exome with a major copy number of two or more (MCN2) (P = 0.05; Figure 5b). Previous pan-cancer analyses of approximately 10,000 patients with advanced cancer defined tumors with ≥50% of autosomal regions showing MCN2 as whole-genome doubled (WGD) (28). Using this criterion, 24.4% (10/41) of *EGFR*-mutant tumors were classified as WGD-positive. In contrast, none of the 17 non-*EGFR* CAG–mutant tumors exhibited WGD. WGD frequency differed significantly between groups (P = 0.026, Fisher’s exact test; Figure 5c), suggesting that WGD is a secondary evolutionary event, preferentially associated with *EGFR* mutation during lung adenocarcinoma development.

**Figure 5.**
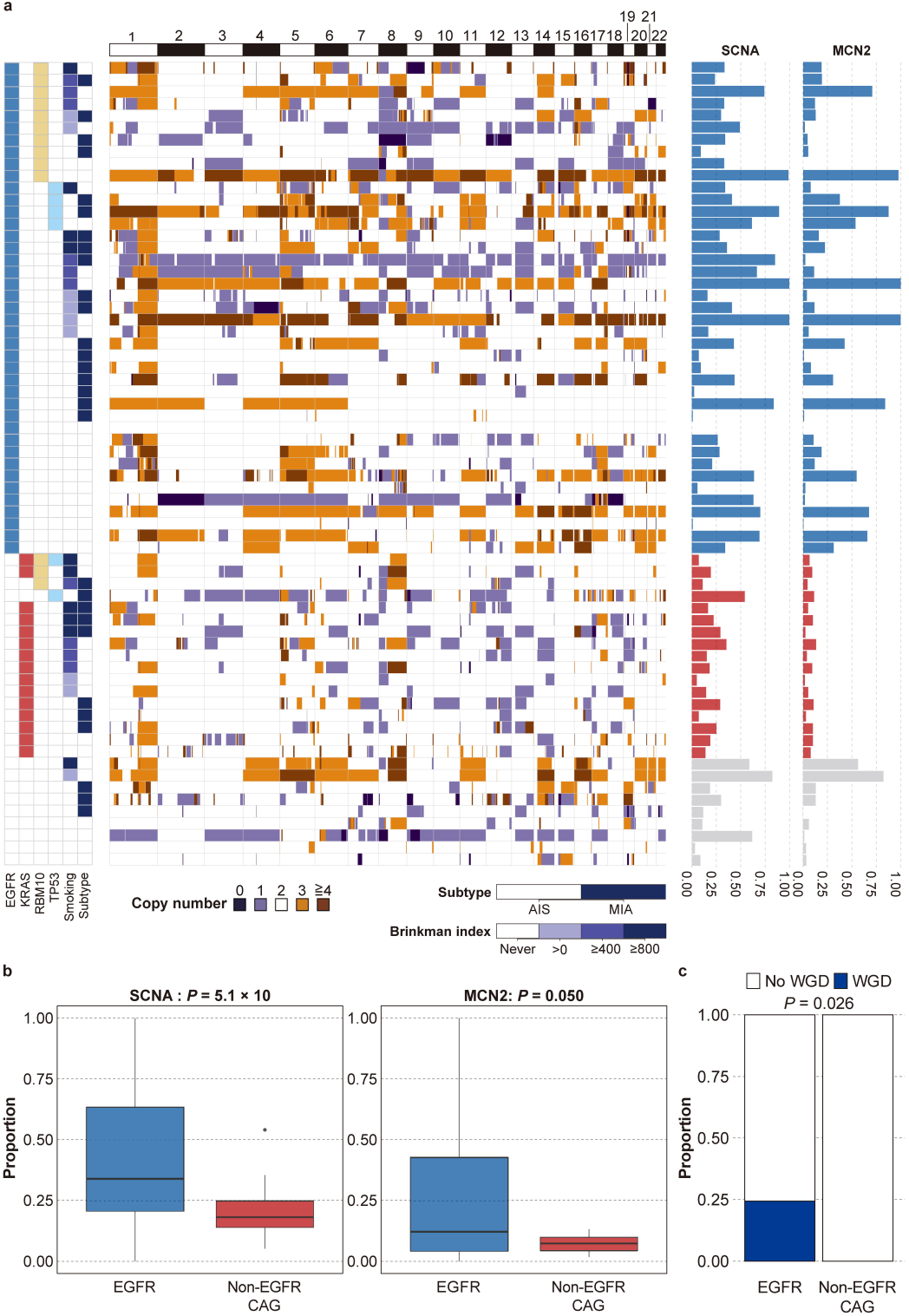
Somatic copy number alterations and whole-genome doubling. (A) Box plots showing the proportion of the exome affected by somatic copy number alterations (SCNAs) in *EGFR*-mutant tumors (n = 41) versus non-*EGFR* CAG–mutant tumors (n = 17). CAG represents mutations in the cancer-associated genes *KRAS*, *LRP1B*, *NF1*, *MET*, *ERBB2*, *STK11*, and *BRAF*. P-values were determined using the exact Wilcoxon rank-sum test. (B) Box plots showing the proportion of the exome with a major copy number (MCN) ≥2. (C) Frequency of whole-genome doubling (WGD), defined as ≥50% of the autosomal genome with MCN ≥2, stratified by driver mutation status. P-values were calculated using Fisher’s exact test.

### Multiregional sequencing of early lung adenocarcinoma

We performed multiregional sequencing in 12 early-stage lung adenocarcinoma cases, each containing three *in situ* and/or invasive lesions plus one adjacent histologically normal-appearing alveolar tissue sample (Figure 6, Supplementary Figures 5-1,2, and 3). Eleven of the 13 tumors included both *in situ* and invasive regions. Multiple regions from the same tumor shared many somatic mutations, supporting a common clonal origin despite substantial intratumor genomic heterogeneity.

**Figure 6.**
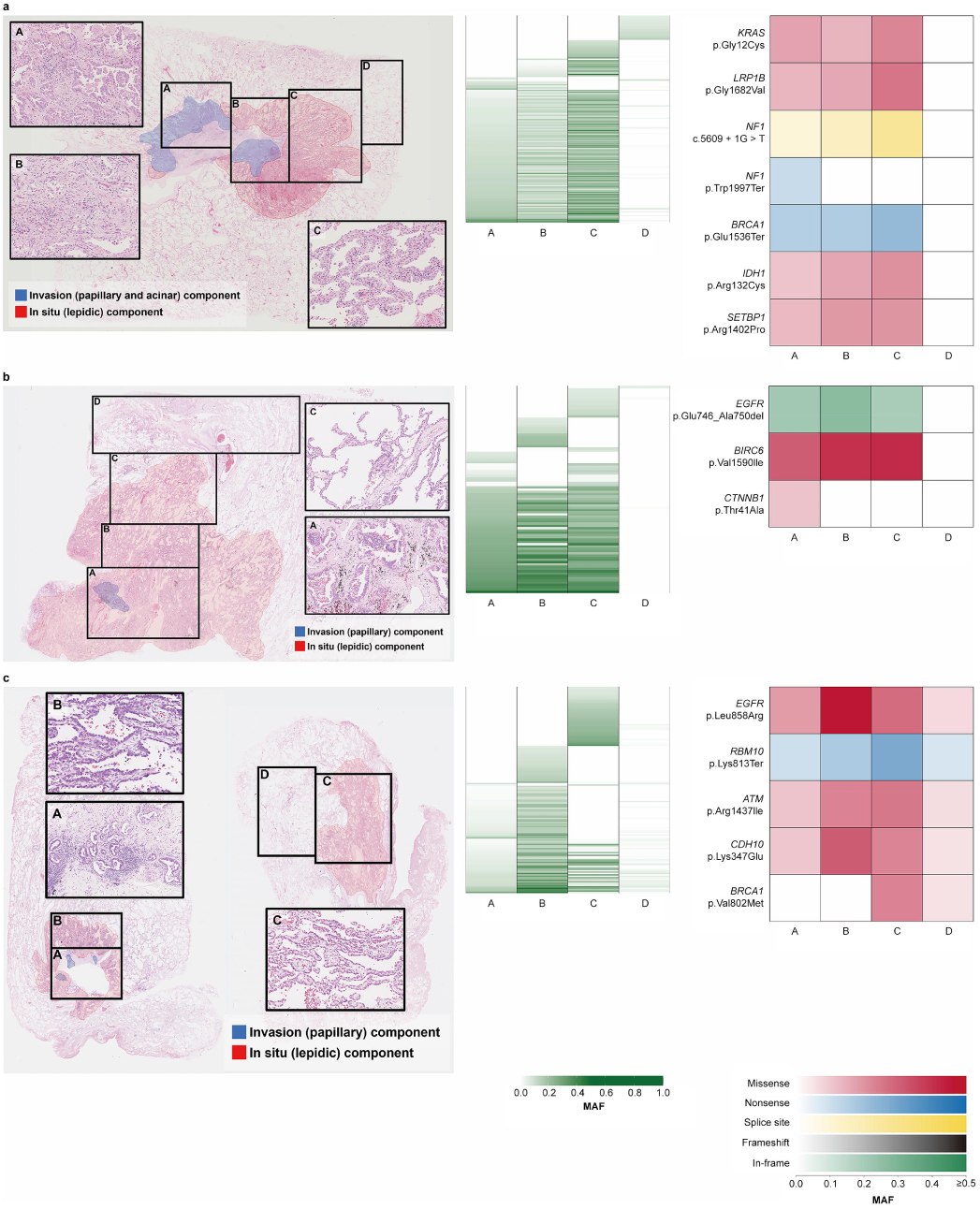
Intratumoral heterogeneity in early-stage lung adenocarcinoma. (A–C) Representative multiregion whole-exome sequencing analyses of early-stage tumors (n = 13). Left: Hematoxylin and eosin–stained sections showing histologically distinct regions (A–C) and adjacent normal-appearing alveolar tissue (D). Red areas indicate noninvasive (*in situ*) components of the adenocarcinoma and blue areas correspond to invasive components. Cases containing blue regions were classified as minimally invasive adenocarcinoma (MIA), whereas cases lacking blue regions were classified as adenocarcinoma *in situ* (AIS). Insets show higher-magnification views area. Right: Heatmaps of mutant allele frequencies (MAFs) for somatic mutations detected in each region. Color intensity corresponds to MAF (scale: 0.0–1.0). (A) G19_26, case with a shared clonal *NF1* splice-site mutation with a region-specific *NF1* nonsense mutation. (B) G19_39, case sharing an *EGFR* driver mutation with a region-specific *CTNNB1* mutation. (C) G22_67, serial sections showing spatially separated regions (A and B vs. C) with distinct mutational profiles; region D represents histologically normal tissue sharing mutations with region C.

One early lung adenocarcinoma containing both *in situ* and invasive regions shared a splice-site mutation in neurofibromin 1 (*NF1*; c.5609 + 1G > T), whereas only the invasive region harbored an additional nonsense *NF1* mutation (p. W1997X), resulting in biallelic of loss of this tumor suppressor gene (Figure 6A). In another early-stage lung adenocarcinoma case, one invasive and two *in situ* regions shared an *EGFR* driver mutation (p. E746_A750del), whereas only the invasive region harbored a missense mutation in catenin beta 1 (*CTNNB1*; p. T41A), a well-known oncogenic mutation in the N-terminal region of the β-catenin protein (Figure 6B). Analysis of spatially separated regions within the same tumor (Figure 6C, showing serial sections of the same tumor used for macrodissection) revealed that closely-located invasive and *in situ* regions (A and B) shared similar mutation profiles, whereas a distantly located *in situ* region (C) exhibited a distinct profile. Histologically normal-appearing alveolar tissue adjacent to region C also shared somatic mutations with that region, supporting the field cancerization concept.

Across all cases, invasive and *in situ* regions within the same tumor consistently shared driver mutations in *EGFR*, *KRAS*, and *MET*, indicating that these alterations arise early during tumorigenesis. Collectively, these findings suggest that secondary cancer-associated gene mutations, along with epigenetic and immune microenvironmental changes, contribute to acquisition of invasive phenotypes in lung adenocarcinoma.

## DISCUSSION

In this study, we analyzed early lung adenocarcinoma cases (38 AIS, 29 MIA) using WES to define the mutational landscape of precursor lesions. A prior large-cohort WGS study on LCIN, composed primarily of adenocarcinomas, reported markedly different driver mutation frequencies in the receptor tyrosine kinase–Ras pathway, particularly for *EGFR* (10). *EGFR* mutations occurred in 61.2% of cases in our cohort compared with 30.6% in the LCIN cohort, whereas mutation frequencies for *KRAS* (9.0% vs. 7.3%), *MET* (4.4% vs. 4.3%), and *ERBB2* (3.0% vs. 3.9%) were similar between studies. The disparity in *EGFR* mutations was even more pronounced when restricting analysis to the never-smoker group in our cohort, reaching a rate of 67.6% (25/37 cases). Because receptor tyrosine kinase–Ras pathway mutations are typically mutually exclusive and persist throughout progression from early to invasive lung adenocarcinoma, this difference likely reflects the distinct ancestral backgrounds of the cohorts: our cohort was exclusively East Asian (Japanese), whereas the LCIN cohort comprised mainly Europeans and Africans. Our findings are also consistent with a recent Japanese WGS study on early lung adenocarcinoma reporting *EGFR* mutation rates of 65.4% for AIS and 66.6% for MIA (29). Together, these observations underscore the urgent need for genomic studies focusing on early lung adenocarcinoma in non-Asian populations to clarify ancestry-specific mechanisms underlying tumorigenesis in these groups.

To determine the timing of driver gene mutations during lung adenocarcinoma development, we compared our data with the IntOGen database, which primarily contains advanced lung adenocarcinoma data. In this comparison, *EGFR* and *RBM10* mutations were more frequent in early-stage lung adenocarcinoma, whereas *TP53*, *KRAS*, and *KEAP1* mutations were less common. Although such comparisons may be confounded by genetic ancestry and smoking differences between cohorts, the results suggest that cancer-associated genes exert differential effects between the early and invasive disease stages. Therefore, future analyses of tumor evolution should incorporate cohort stratification by ancestry and smoking history.

We also examined associations between genetic mutations and clinicopathological or lifestyle factors. Consistent with previous reports, we found that *KRAS* mutations were overrepresented in moderate-to-heavy smokers (30). Additionally, mutations in *LRP1B* and *RBM10* showed nominal associations with moderate-to-heavy smoking. Although the *KRAS*–smoking relationship is well established, links between tobacco exposure and *LRP1B* or *RBM10* mutations have rarely been reported in lung adenocarcinoma (31,32). This observation may reflect our precise quantification of smoking exposure, allowing trend detection. In contrast, lower BMI, previously associated with lung cancer risk (33), potentially due to the reduced adipose tissue capacity for storing and metabolizing carcinogen–DNA adducts, showed no significant association with gene mutation status or mutational burden in early lung adenocarcinoma. These findings align with larger Japanese epidemiological studies investigating relationships between genomic mutations and lifestyle factors, including BMI (30).

Patterns of co-occurrence and mutual exclusivity among cancer-associated gene mutations were already evident during the early stages of lung adenocarcinoma development. *RBM10* mutations occurred frequently (19.4%) in our cohort. Located on the X chromosome, *RBM10* regulates alternative splicing of multiple target genes (21), and functional studies have shown that *RBM10* disruption contributes to lung adenocarcinoma pathogenesis through splicing deregulation (34). In our analysis, *RBM10* mutations represented a broad range of inactivating alterations, supporting its tumor-suppressive role in early lung adenocarcinoma. Notably, *RBM10* and *EGFR* mutations co-occurred in 24.4% (10/41) of early lung adenocarcinoma cases. In mouse models of *Egfr*-driven, *Tp53*-deficient lung adenocarcinoma, *Rbm10* inactivation strongly promotes tumor growth (35). However, we did not detect concurrent *RBM10* and *TP53* mutations in *EGFR*-mutant early tumors. This finding suggests that highly aggressive tumor growth driven by this combinatorial mutation profile may emerge in advanced-stage lung adenocarcinoma when *TP53* mutation rates are higher. These results support early detection and surgical resection of lung adenocarcinoma before accumulation of high-risk genetic combinations, such as concurrent *RBM10* and *TP53* mutations, potentially aided by polygenic risk stratification (36).

Tumor mutational burden in our cohort was driven largely by C > A substitutions, a signature associated with tobacco exposure. To further characterize mutational processes, we stratified cases into five groups: *EGFR*-NS, *EGFR*-SM, non-*EGFR* CAG-NS, non-*EGFR* CAG-SM, and No CAG-NS. The *EGFR*-NS group showed significant enrichment of SBS2 and SBS13, indicating aberrant APOBEC3 activation (37). Although viral infection may trigger APOBEC3 activation, evidence linking viruses, such as human papillomavirus, to *EGFR*-NS lung adenocarcinoma remains inconsistent and largely negative (30,38). Clarifying the causes of APOBEC3 activation in this subgroup will require integrated virological and genomic polymorphism analyses, particularly because APOBEC3-related carcinogenesis varies across genetic backgrounds (39). Environmental exposure may also contribute. Prior studies reported enrichment of SBS4 and SBS5 in never-smoker lung cancers associated with air pollution exposure (40), suggesting that some SBS signatures observed in the *EGFR*-NS and non-*EGFR* CAG-NS groups may reflect environmental pollutants, such as PM2.5. Another notable finding was the clock-like mutational pattern in *EGFR*-mutant, but not non-*EGFR*-mutant, tumors. A previous WGS study of 346 never-smoker lung adenocarcinoma cases similarly showed enrichment of SBS5/SBS40a signatures in *EGFR*-mutated tumors, attributed mainly to aging or other endogenous processes, and reported longer latency in *EGFR*-mutant tumors compared with wild types (41). Taken together, these findings support an age-related evolutionary pathway for *EGFR*-mutant lung adenocarcinoma that establishes early in tumorigenesis.

*EGFR*-mutant lung adenocarcinoma occurs predominantly in Asian nonsmoking females but is also common in smokers. In our cohort, 16 of 41 (39.9%) *EGFR*-mutant cases occurred in smokers (*EGFR*-SM). Although smokers exhibited higher C > A mutational burdens compared with nonsmokers in *EGFR*- and non-*EGFR* CAG–mutant groups, the increase was significantly elevated in the non-*EGFR* CAG-SM group. This discrepancy may reflect different cellular origins. Prior studies demonstrated substantial variation in mutational burden among colonies derived from single bronchial epithelial cells in smokers (42). Pathological studies have also distinguished terminal respiratory unit (TRU)-type and non-TRU-type lung adenocarcinoma (43). Collectively, these findings suggest that *EGFR*-SM tumors arise from TRU cells, which may accumulate relatively few mutations, whereas non-*EGFR* CAG-SM tumors derive from non-TRU cells that are more susceptible to mutation accumulation. As shown in Figure 3c, strong transcription strand asymmetry of SBS4-related C > A mutations was observed in non-*EGFR* CAG-NS and non-*EGFR* CAG-SM tumors. Tobacco carcinogens, such as benzo[a]pyrene, predominantly form DNA adducts on guanine bases (44). Transcription-coupled nucleotide excision repair (TC-NER) efficiently removes lesions from the transcribed strand (45), whereas damage persists on the nontranscribed strand, producing excess G > T mutations on this strand. Because somatic mutations are conventionally represented by pyrimidine-base substitutions, these G > T mutations are reported as complementary C > A mutations on the transcribed strand. We also observed transcriptional strand bias for SBS5-associated T > C mutations, enriched on the transcribed strand. Although the etiology of SBS5 remains unclear (26), TC-NER–mediated removal of adenine damage from the transcribed strand may explain this strand asymmetry (45).

WGD promotes genomic instability and malignant progression in cancer because WGD-positive cells are susceptible to replication stress–induced DNA damage (46). WGS-based studies have indicated that WGD occurs in approximately 35% of lung adenocarcinomas and over 50% of *TP53*-mutant non-small cell lung cancers (28). However, as these data are derived mainly from advanced tumors, the clinical significance of WGD in early lung adenocarcinoma remained uncertain. In the present study, nearly one-quarter of *EGFR*-mutant early lung adenocarcinomas showed evidence of WGD, whereas none of the non-*EGFR* CAG–mutant tumors exhibited WGD. This finding aligns with a previous report showing frequent chromosomal numerical abnormalities even in early-stage lung adenocarcinoma (47). Together, these results suggest that WGD contributes early to the development of *EGFR*-mutant tumors. Computational models indicate that hyperactive EGFR signaling and downstream phosphatidylinositol-3-kinase/protein kinase B pathway activation can dysregulate polo-like kinase 1 expression, promoting defective cytokinesis and chromosome segregation that lead to aneuploidy (48). Moreover, the frequent coexistence of WGD with *EGFR* and/or *TP53* mutations in early lung adenocarcinoma further supports the rationale for early detection and resection, as tumors harboring these combined abnormalities may be predisposed to tyrosine kinase inhibitor resistance (49).

This study is limited by its relatively small cohort of 67 Japanese patients and the use of whole-exome sequencing rather than whole-genome sequencing. Despite these limitations, our results define two distinct evolutionary trajectories in early lung adenocarcinoma: an age-related pathway featuring *EGFR*-linked genome doubling and a smoking-driven pathway. We demonstrate that genomic divergence begins at the preinvasive stage, with key driver mutations acting as shared trunk events across *in situ* and invasive regions. These insights highlight the critical role of early whole-genome doubling in *EGFR*-mutant cases.

## Data Availability

All data produced in the present study are available upon reasonable request to the authors

## Acknowledgements

The authors would like to thank Prof. Daichi Maeda (Department of Molecular and Genomic Pathology, Kobe University) and Dr. Hirofumi Rokutan (Department of Pathology, Tokyo Metropolitan Institute of Gerontology) for their helpful discussions regarding this study. We also express our gratitude to Mr. Yukitsugu Kudo (Department of Cellular and Organ Pathology, Akita University), Junko Kajiwara, Junko Kitayama, and Yumiko Sato (Human Genetics Laboratory, National Institute of Genetics) for their excellent technical support.

## Financial Support

This work was supported by JSPS KAKENHI (Grant Nos. 19H03446 to Y.I., 23K27447 to H.N., and 24K10104 to A.G.), a Challenging Exploratory Research Projects for the Future grant from the Research Organization of Information and Systems (ROIS) (H.N.), and the SRF (A.G. and H.S.).

**Supplementary Figure 1. Distribution of driver mutations in *KRAS* and *MET*.** (A) Lollipop plot of *KRAS* mutations (n = 6) identified in the cohort. (B) Lollipop plot of *MET* mutations (n = 3). Mutations are color-coded by type (missense or splice site). Protein domains are labeled; P-loop, phosphate-binding loop.

**Supplementary Figure 2. Associations between mutational burden and clinicopathologic features.** (A–G) Box plots showing somatic mutation burdens per megabase for indicated substitution types and signatures (n = 67). Center lines indicate medians, boxes represent interquartile ranges, and whiskers extend to 1.5× the interquartile range. 1 (A) Comparison between adenocarcinoma *in situ* (AIS; n = 38) and minimally invasive adenocarcinoma (MIA; n = 29). 1 (B) Comparison by patient sex. 2 (C) Association with patient age (decades). 2 (D) Comparison by BMI (obese ≥25; nonobese <25). 2 (E) Stratification by cumulative smoking exposure (SI). 2 (F) Comparison by drinking history (yes vs. no). 2 (G) Comparison by driver gene mutation status (*EGFR* vs. non-*EGFR* CAG vs. no CAG). P-values were calculated using the Mann–Whitney U test (two groups) or Kruskal–Wallis test (three or more groups).

**Supplementary Figure 3. Interaction effect between smoking exposure and driver gene mutation status on T > C mutation burden.** (A–D) Comparisons of T > C mutation burden between nonsmokers and smokers (A) or according to cumulative smoking exposure measured via the SI (C), stratified by driver gene mutation status. Coefficient plots from multivariable linear regression models show independent and interaction effects of smoking and driver gene mutation status on T > C mutation burden, with smoking modeled as a binary variable (nonsmoker vs. smoker) (B) or cumulative exposure via the Brinkman index (D).

**Supplementary Figure 4. Interaction effect between smoking exposure and driver gene mutation status on total mutation burden.** (A–D) Comparisons of total mutation burden between nonsmokers and smokers (A) or according to cumulative smoking exposure measured via the SI (C), stratified by driver gene mutation status. Coefficient plots from multivariable linear regression models show independent and interaction effects of smoking and driver gene mutation status on total mutation burden, with smoking modeled as a binary variable (nonsmoker vs. smoker) (B) or cumulative exposure measured via the SI (D).

**Supplementary Figures 5-1,2, and 3. Intratumoral heterogeneity in early-stage lung adenocarcinoma.** Multiregion whole-exome sequencing of early-stage lung adenocarcinomas. Upper panels: Hematoxylin and eosin–stained sections showing histologically distinct regions (A–C) and adjacent normal-appearing alveolar tissue (D). Red areas correspond to noninvasive (*in situ*) components of the adenocarcinoma, whereas blue areas indicate invasive components. Cases containing blue regions were classified as minimally invasive adenocarcinoma (MIA), whereas cases lacking blue regions were classified as adenocarcinoma *in situ* (AIS). Insets show higher-magnification views of each area. Lower panels: Heatmaps of mutant allele frequencies (MAFs) for somatic mutations identified in each region. Color intensity corresponds to MAF (scale: 0.0–1.0).

**Supplementary Table 1. Characteristics of patients.**

**Supplementary Table 2. Average sequencing depth and coverage of WES for each sample.**

**Supplementary Table 3. Associations between clinicopathological features and gene mutation status.**

**Supplementary Table 4. Comparison of somatic mutation frequencies in 76 genes between early lung adenocarcinoma (current study) and invasive lung adenocarcinoma (IntOGen database).**

**Supplementary Table 5. Associations of somatic mutation burdens with clinicopathological features and gene mutation status**.

