## Supplementary material for "Distinct Evolutionary Trajectories in Early Lung Adenocarcinoma: Age-Related Pathway with Epidermal Growth Factor Receptor–Associated Genome Doubling and Smoking-Driven Pathway": Suppl Notes

**Supplementary notes**

**Quality control of sequencing data**

Sequencing data quality control was performed as described previously [ref 1]. Briefly, the Illumina adapter sequences were trimmed using Trim Galore (v0.6.3; (<https://www.bioinformatics.babraham.ac.uk/projects/trim_galore/>). Low-quality reads were removed or trimmed with Trimmomatic (v0.39) [ref 2]. Filtered reads were aligned to the human reference genome (GRCh38), including sequence decoys and viral sequences generated by the Genomic Data Commons of the National Cancer Institute, using BWA-MEM (v0.7.17) [refs 3 and 4] (arXiv:1303.3997). Sequence alignment map files were sorted and converted to binary alignment map (BAM) format via SAMtools (v1.9) (arXiv:1303.3997) [ref 4]. BAM files were processed using Picard tools (v2.20.6) (<http://broadinstitute.github.io/picard/>) to remove PCR duplicates. Base-quality recalibration was conducted using GATK (v4.1.3.0) [refs 5 and 6]. Average sequencing depth and target-region coverage were calculated with SAMtools. BEDOPS (v2.4.36) [ref 7] and BEDTools (v2.28.0) [ref 8] were used to process FASTA, VCF, and BED files.

**Variant calling**

Somatic SNVs and short indels were identified in paired lung adenocarcinoma and matched background lung tissues using Strelka2 (v2.9.10) [44]. Somatic indel calling incorporated candidate indel sites detected via Manta (v1.6.0) [45]. Variants with empirical scores >13.0103 (= −10 × log10 0.05) from Strelka2 were retained for downstream analyses. To reduce false-positive calls, we excluded variants with frequencies ≥0.001 in any reference population from the 1000 Genomes Project [46], the National Heart, Lung, and Blood Institute GO Exome Sequencing Project [47], or the Genome Aggregation Database [48]. High-confidence somatic variants met the following criteria: (i) sequencing depth ≥20; (ii) ≥8 reads supporting the mutant allele in tumor tissue; (iii) mutant allele frequency (MAF) ≤0.01 in matched background lung tissue; and (iv) <2 mutant-supporting reads in matched background tissue. To further improve call accuracy, we compiled MAF profiles for candidate variants across all control samples by counting reference and mutant reads with SAMtools mpileup [49]. Only reads with mapping quality >30 and base quality >20 were included. Variants with MAF ≥0.01 and multiple mutant-supporting reads in ≥2 background lung samples were excluded. Functional annotation of protein-coding and transcription-related effects was performed using Ensembl Variant Effect Predictor [50]. Curated information on cancer-associated genes and their functional roles was retrieved from the COSMIC database [51].

**Visualizations**

Lollipop plots showing mutation distributions and protein domains were generated using the trackViewer R package [ref 9]. Circular plots illustrating co-occurrence and mutual exclusivity of cancer-associated gene mutations were generated using the circlize R package [ref 10].
