## Supplementary material for "Distinct Evolutionary Trajectories in Early Lung Adenocarcinoma: Age-Related Pathway with Epidermal Growth Factor Receptor–Associated Genome Doubling and Smoking-Driven Pathway": Suppl Table 1

Supplementary Table 1. Patient characteristics.

| **Characteristics** | **AIS (n = 38)** | **MIA (n = 29)** |
| --- | --- | --- |
| Age |  |  |
| Median (range), year | 70.5 (50–83) | 72 (58–81) |
| Distribution, count (%) |  |  |
| ≥50 years | 5 (13.2) | 1 (3.4) |
| ≥60 years | 11 (28.9) | 11 (37.9) |
| ≥70 years | 15 (39.5) | 16 (55.2) |
| ≥80 years | 7 (18.4) | 1 (3.4) |
| Sex, count (%) |  |  |
| Male | 18 (47.4) | 11 (37.9) |
| Female | 20 (52.6) | 18 (62.1) |
| Body-mass index |  |  |
| Median (range), kg/m^2^ | 22.4 (17.8–29.2) | 23.6 (15.5–40.0) |
| Distribution, count (%) |  |  |
| Underweight (≤18.5) | 1 (2.6) | 2 (6.9) |
| Normal (>18.5) | 30 (78.9) | 18 (62.1) |
| Obese (>25.0) | 7 (18.4) | 9 (31.0) |
| Smoking index^a^, count (%) |  |  |
| Nonsmoker (= 0) | 20 (52.6) | 18 (62.1) |
| Mild (>0) | 6 (15.8) | 3 (10.3) |
| Moderate (≥400) | 7 (18.4) | 3 (10.3) |
| Heavy (≥800) | 5 (13.2) | 5 (17.2) |
| Alcohol drinking status, count (%) |  |  |
| Nondrinker | 20 (52.6) | 18 (62.1) |
| Drinker | 15 (39.5) | 11 (37.9) |
| Missing | 3 (7.9) | 0 (0.0) |

^a^ Smoking index is calculated as cigarettes smoked per day × years of smoking.
