## Supplementary figures and images for "Distinct Evolutionary Trajectories in Early Lung Adenocarcinoma: Age-Related Pathway with Epidermal Growth Factor Receptor–Associated Genome Doubling and Smoking-Driven Pathway"

### Fig S1

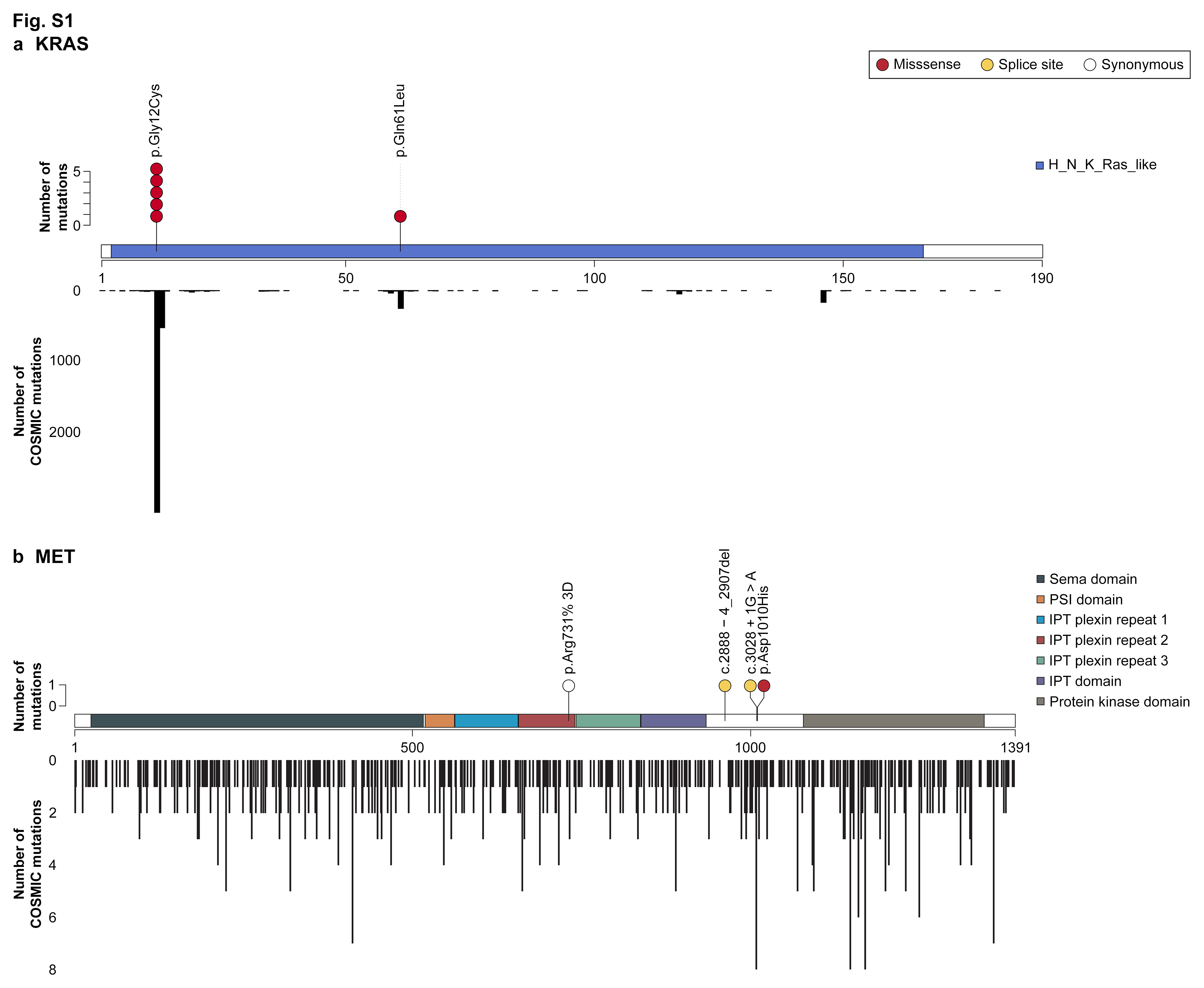

### Fig S2 1

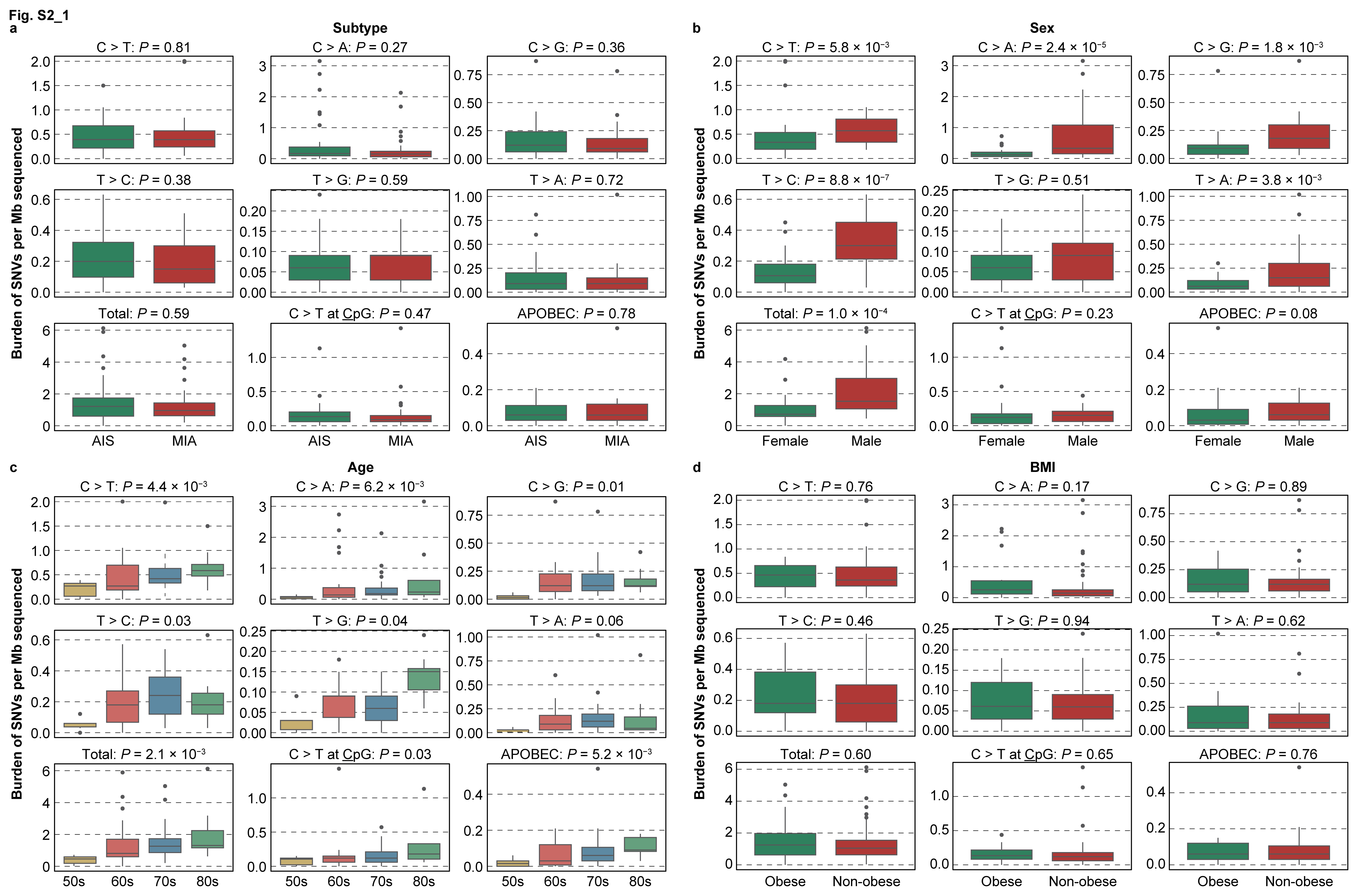

### Fig S2 2

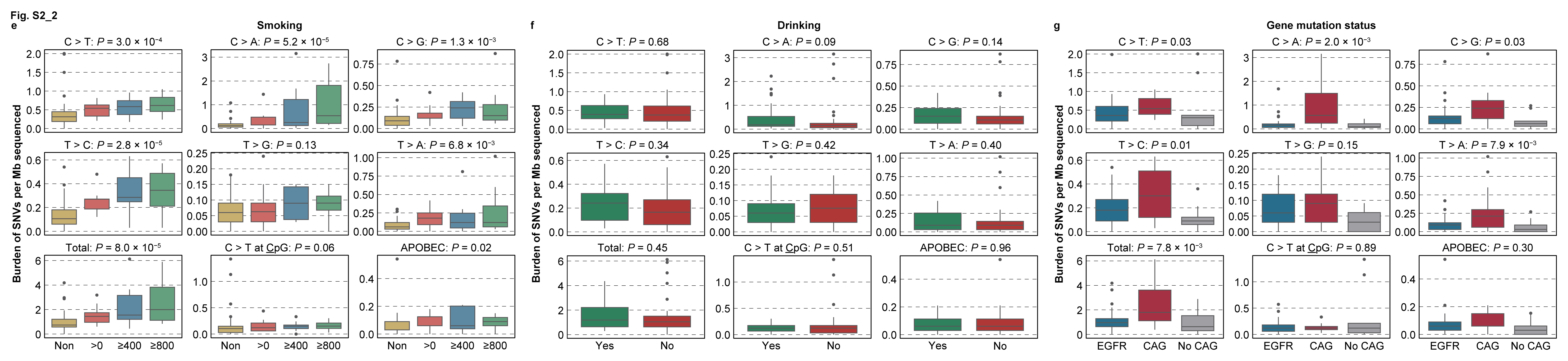

### Fig S3

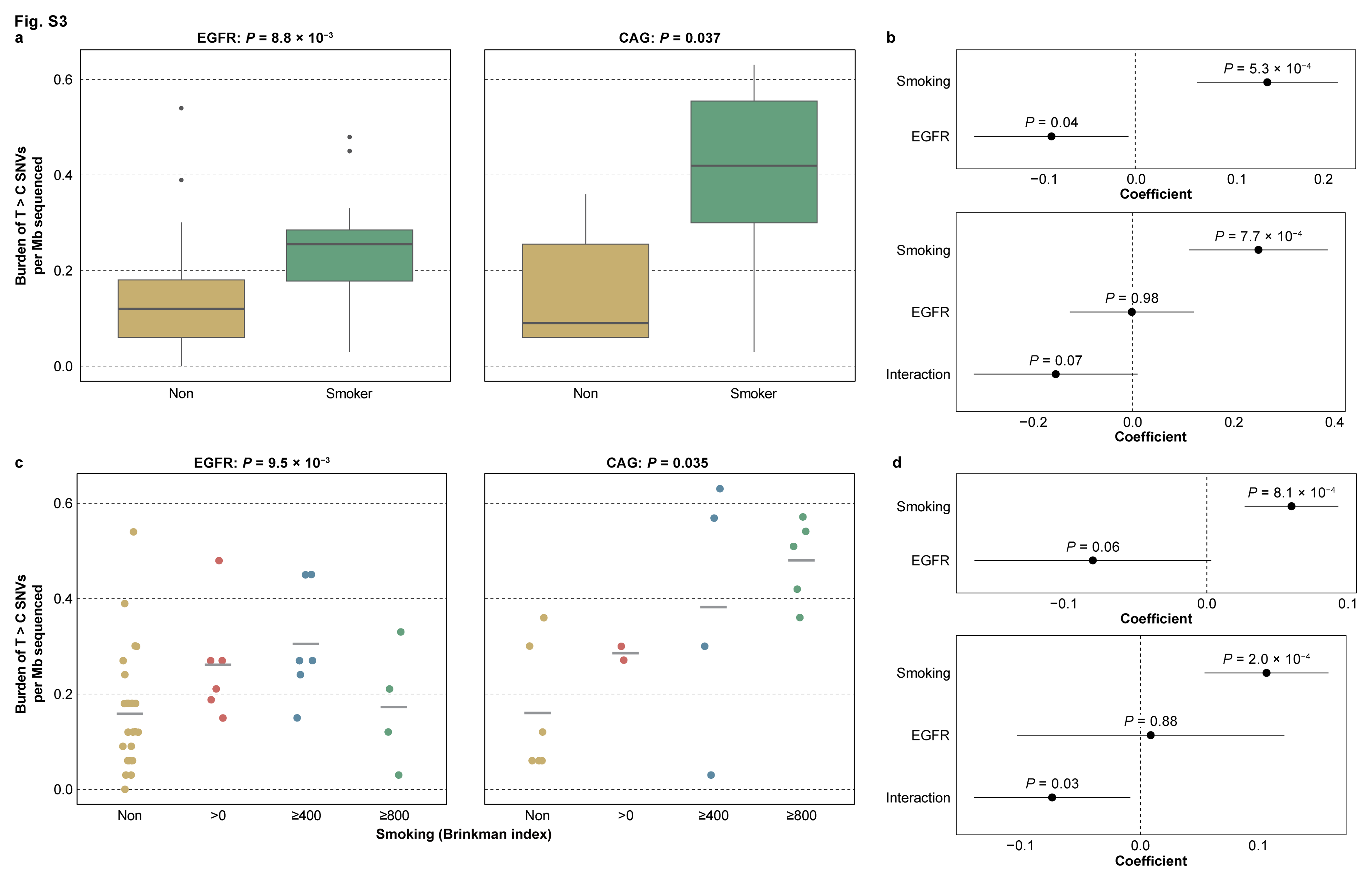

### Fig S4

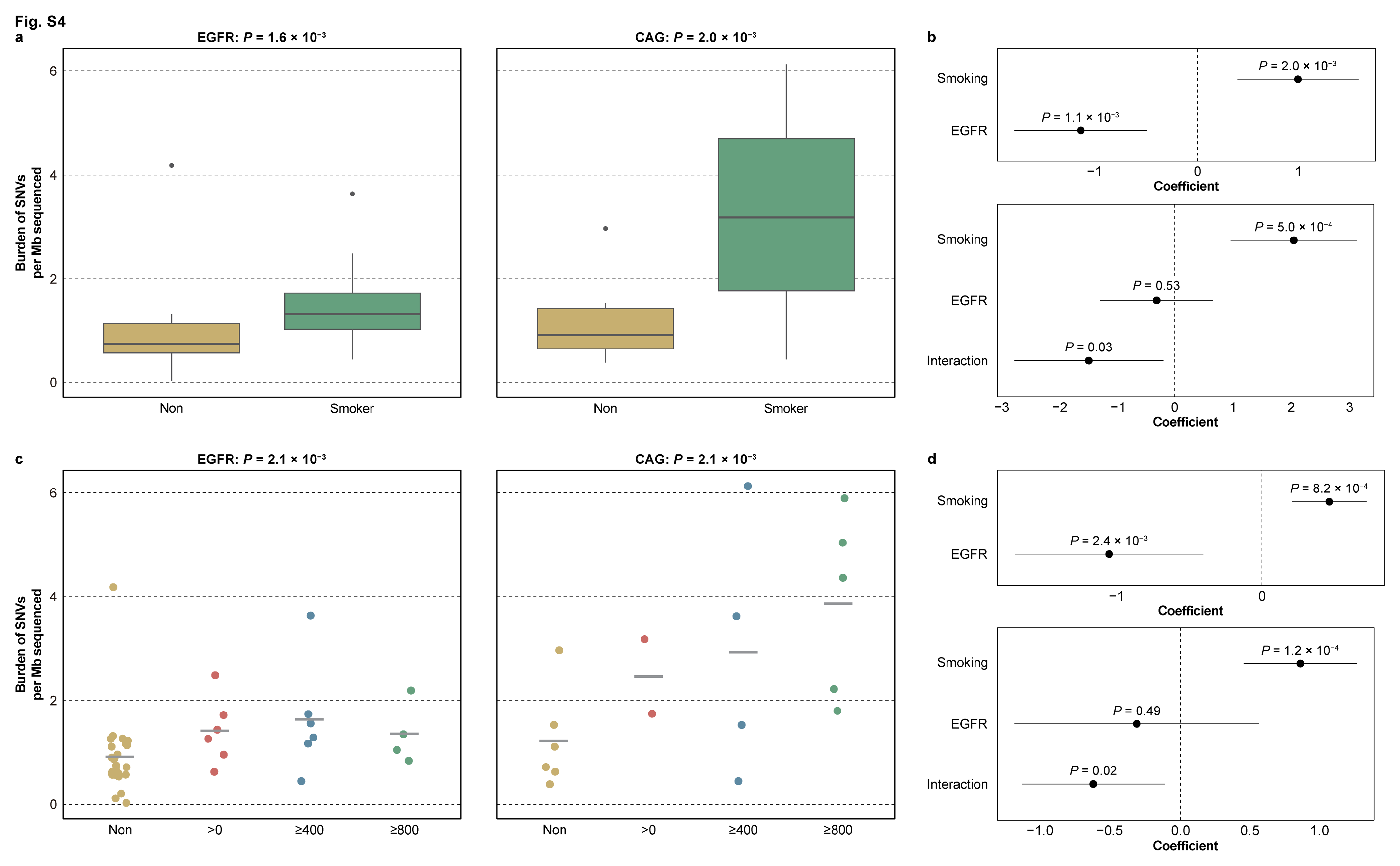

### Fig S5 1

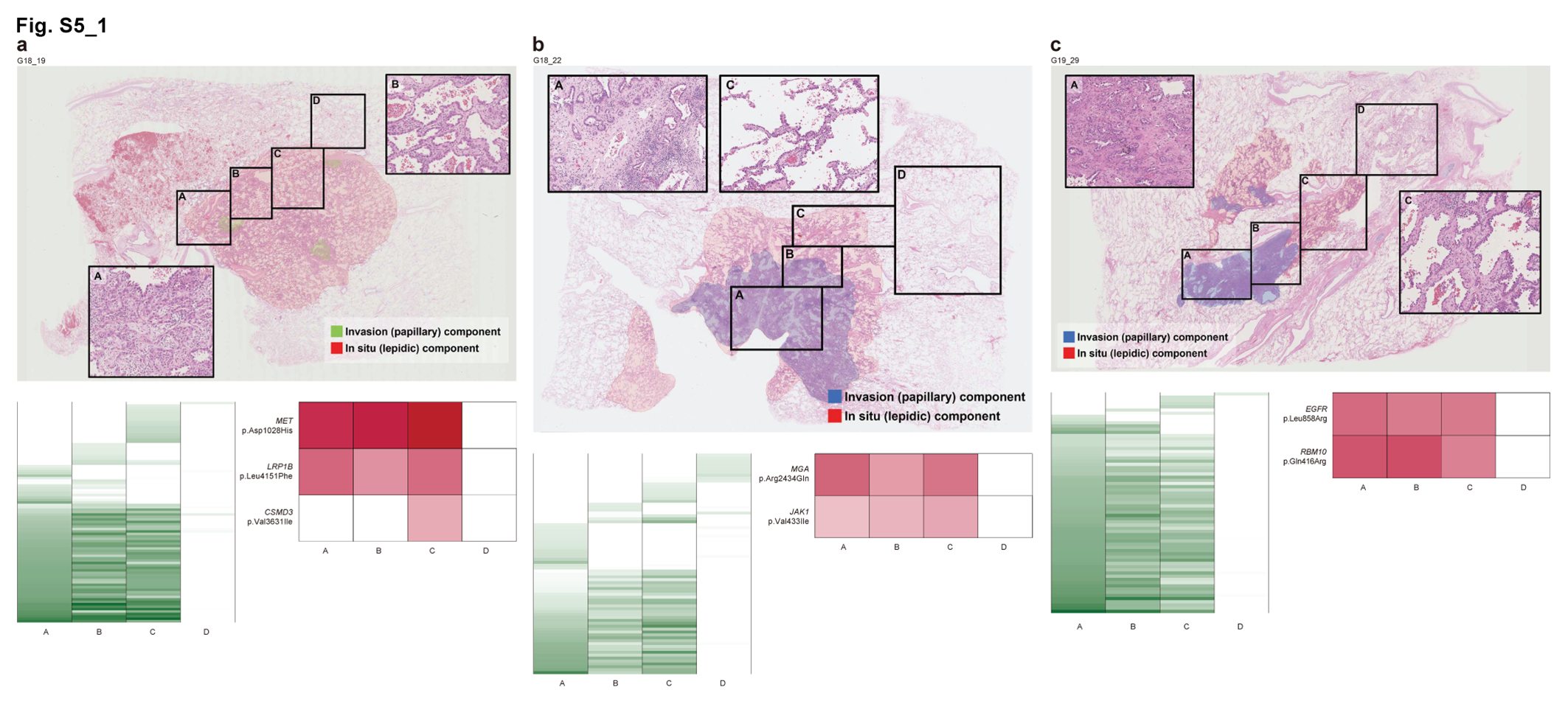

### Fig S5 2

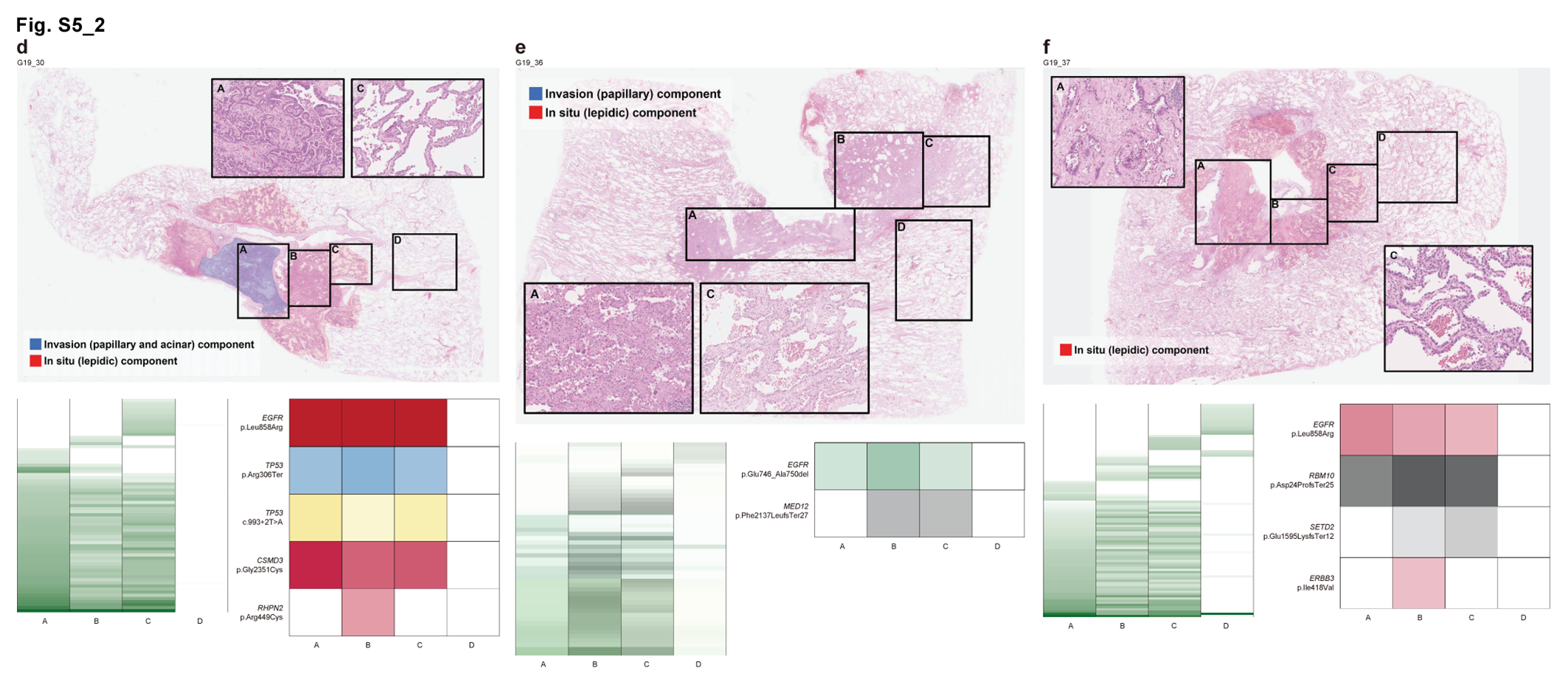

### Fig S5 3

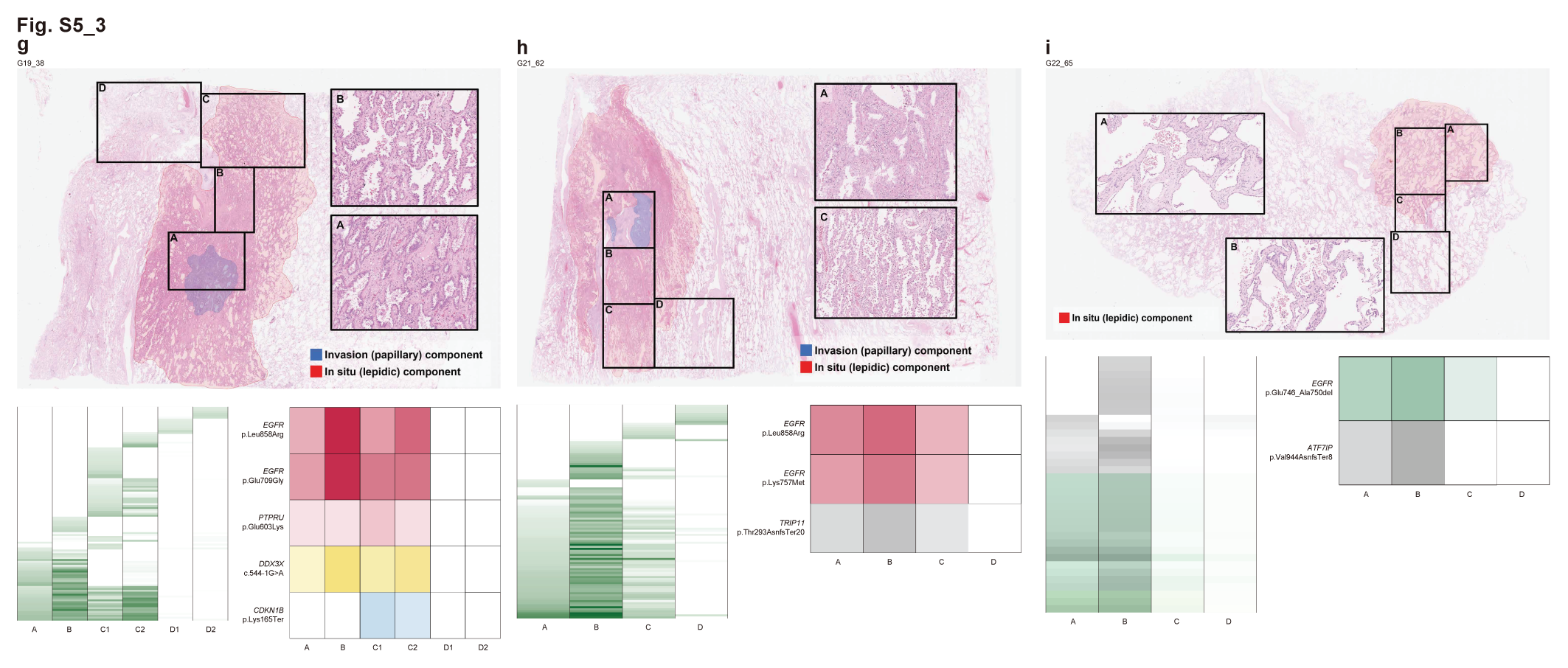
